# Contrasting methods of measurement of antibiotic exposure in clinical research: a real-world application predicting hospital-associated *Clostridioides difficile* infection

**DOI:** 10.1101/2024.01.15.24301334

**Authors:** Jessica L. Webster, Stephen Eppes, Brian K Lee, Nicole S. Harrington, Neal D. Goldstein

## Abstract

The goal of this article is to summarize common methods of antibiotic measurement used in clinical research and demonstrate analytic methods for selection of exposure variables. Variable selection was demonstrated using three methods for modeling exposure, using data from a case-control study on *Clostridioides difficile* infection in hospitalized patients: 1) factor analysis of mixed data, 2) multiple logistic regression models, and 3) Least Absolute Shrinkage and Selection Operator (LASSO) regression. The factor analysis identified 9 variables contributing the most variation in the dataset: *any antibiotic treatment*; *number of classes*; *number of treatments*; *dose*; and classes *monobactam*, β*-lactam* β*-lactamase inhibitors*, *rifamycin*, *carbapenem*, and *cephalosporin*. The regression models resulting in the best model fit used predictors *any antibiotic exposure* and *proportion of hospitalization on antibiotics*. The LASSO model selected 22 variables for inclusion in the predictive model, exposure variables including: *any antibiotic treatment*; classes β*-lactam* β*-lactamase inhibitors*, *carbapenem*, *cephalosporin*, *fluoroquinolone*, *monobactam*, *rifamycin*, *sulfonamides*, and *miscellaneous*; and *proportion of hospitalization on antibiotics*. Investigators studying antibiotic exposure should consider multiple aspects of treatment informed by their research question and the theory on how antibiotics may impact the distribution of the outcome in their target population.

## INTRODUCTION

The study of antibiotic exposure in clinical research is common in studies evaluating the impact of antibiotics on hospital-associated infections (such as *Clostridioides difficile*),^1–7^ the human microbiome,^8,9^ immune-mediated disorders,^10–14^ among others.^15–17^ It is equally important for health systems to quantify antibiotic administration in an effort to reduce unnecessary use through antimicrobial stewardship programs.^18–22^ Investigators involved in such research are often faced with the challenge of determining how to measure and define (i.e., operationalize) antibiotic exposure, either in individual patients or aggregated to a larger scale, such as a hospital or a health system. In the most simplistic sense, antibiotic treatment can be operationalized as a dichotomous treatment, capturing whether a patient received any antibiotic over the course of follow-up. Yet such simplification ignores the reality that antibiotic treatment can be a complex exposure that changes over time, and variations in treatment characteristics may have differential effects on the outcome of interest.

### Objectives

In this paper, we aim to summarize, contrast, and demonstrate methods of measuring antibiotic exposure in clinical and epidemiological research. By exposure, we are referring to the salient aspects of treatment that may be modeled as one or more variables in the data. Relevant dimensions of exposure may include type of medication, treatment dose, length of treatment, route of administration, and timing of treatment over follow-up – all of which could vary between patients as well as within patients prescribed multiple courses. We demonstrate three analytic approaches for modeling data with complex antibiotic exposure structures: the first as an exploratory tool (ignoring the outcome of interest), and the remaining as tools to predict hospital-associated *C. difficile* infection. Investigators considering research that involves antibiotics can use this information as a resource in the study design phase when considering the level of detail needed to measure exposure, in the analytic phase when building a predictive model or calculating measures of effect, and in the dissemination phase when comparing results to other studies and identifying potential limitations to the measure(s) and analytic method(s) selected.

## METHODS

Informed by the constructs identified within a methodological rapid review (details and results of which are provided in Supplemental Document 1 and Supplemental Table 1, respectively), we demonstrated three analytic methods for exposure variable exploration and selection using data from a case-control study designed to investigate healthcare-associated *C. difficile* infection.

### Study population

The study was conducted at Hahnemann University Hospital in Philadelphia, Pennsylvania. Data were extracted from electronic medical records (EMR) from August 1^st^, 2014, through May 1^st^, 2018, and included information on *C. difficile* infection, current and prior medication (including antibiotic) use, patient demographics, and comorbidities. Cases were patients aged 18 years or older, who showed symptoms of *C. difficile* infection at least 72 hours after admission to the hospital, and whose infection was confirmed by a rapid enzyme immunoassay test. Controls were patients 18 years or older, who had a length of stay longer than 72 hours and did not have a positive *C. difficile* test between admission and discharge. Additional details related to the study design and data collection are described elsewhere.^23^ A total of 222 cases and 455 controls were selected into the study. Institutional review board approval (no. 1403002707) for this project was granted by Drexel University Human Research Protections Office.

### Antibiotic exposure classifications

Antibiotic exposure was determined through data extracted from the EMR. Up to four unique antibiotic courses were recorded for each patient, with information on the type of medication (name of formulation), treatment dose, and start and stop dates recorded for each unique antibiotic course within the current hospitalization. Seven antibiotic exposure variables were created as summary measures for each patient in the study:

1. any antibiotic exposure,
2. number of unique antibiotic courses,
3. class of antibiotic(s),
4. cumulative dose of antibiotic(s),
5. number of unique antibiotic classes,
6. cumulative length of exposure to antibiotics (days), and
7. the proportion of a patient’s hospitalization on antibiotics (length of exposure divided by length of hospitalization).

Supplemental Table 2 provides a list of antibiotic class variables and the specific medications included within each class.

### Covariates

The following demographic and health characteristics were considered in the predictive models, identified as potential risk factors for *C. difficile* infection and/or associated with the exposure, based on prior research demonstrating the clinical and epidemiological significance: age, sex, race/ethnicity, insurance, referral, body mass index, length of hospitalization (days), Charlson Comorbidity Index, Intensive Care Unit (ICU), non-*C. difficile* infections, procedures, current proton pump inhibitor (PPI) exposure, current steroid exposure, current chemotherapy, prior hospitalizations, prior antibiotics, prior PPI exposure, prior steroid exposure, prior chemotherapy. Two interactions were considered, per existing literature ^24^: current antibiotics × current PPI exposure and prior antibiotics × prior PPI exposure.

### Statistical analysis

The distribution of exposure variables, demographic characteristics, and health covariates were assessed overall and by case-control status. To demonstrate methods of measuring and selecting exposure variable classifications, we used: 1) factor analysis of mixed data, 2) logistic regression, and 3) Least Absolute Shrinkage and Selection Operator (LASSO) regression.

#### Factor analysis of mixed data

We conducted a factor analysis of mixed data to identify the antibiotic exposure variables that together, may better represent the sample population than a single exposure characteristic on its own. In addition, it was suspected that many characteristics of antibiotic exposure may be correlated with each other. Variables were considered strongly correlated if the absolute value of their Pearson correlation coefficient was greater than or equal to 0.70.^25^ Factor analysis of mixed data was performed using the FAMD function within the FactoMineR (Le, Josse, Husson; 2008) R package. Eigenvalues were calculated as the sum of the squared factor scores for each component, and dimensions with an eigenvalue ≥1 were retained based on the Kaiser Criterion.^26^ The contribution of a variable to each dimension was calculated as a ratio of the squared factor scores for that variable by the eigenvalue of the dimension.^27^

#### Logistic regression models

The second approach analyzes separately the association between each operationalization of antibiotic exposure and the outcome, or alternatively develops separate predictive models for each antibiotic exposure variable – this method may be useful when the ideal variable measure is unknown, and/or the researcher would like to predict an outcome based on multiple, separate antibiotic exposure characteristics. We ran seven logistic regression models assessing the probability of *C. difficile* infection based on each antibiotic exposure variable. Demographic characteristics and health covariates whose distributions differed according to the bivariate analyses were adjusted for in each model. Adjusted odds ratios were calculated to estimate the odds of *C. difficile* infection based on levels of each exposure characteristic. Akaike Information Criterion (AIC) was used to assess model fit.

#### Least Absolute Shrinkage and Selection Operator regression

The goal of the final approach was to demonstrate a variable selection method in predictive modeling. The chosen method, LASSO regression, is a machine learning shrinkage and variable selection technique, often used when a dataset contains many potential exposure or predictor variables and the researcher seeks to reduce the number of included variables within a predictive model ^28^. The model was run using the glmnet (Friedman, Tibshirani, Hastie; 2010) R package, which fits generalized linear regression models computed for the LASSO penalty. We first conducted 10-folds cross validation to identify the value of the penalty term (λ) that produced the most predictive model (minimum λ). As a sensitivity analysis we utilized the more conservative value of the penalty term, the maximum value of λ within 1 standard error (SE) of the minimum λ (1SE λ). These values were used to run binomial LASSO models, and variables were ranked by importance according to predicted regression coefficients fitted using the selected model parameters. Predictive models using the variables selected by the two LASSO models were used to calculate coefficients and predictive performance of the models [Area Under the Receiver Operating Characteristic Curve (AUC)] using the caret (Kuhn, M; 2008) R package. We included all covariates and interaction terms as potential predictors.

Data preparation and analyses were conducted using RStudio (R Version 4.2.2, R Core Team, 2022). Continuous variables were centered and scaled, and categorical variables were transformed to numeric using one-hot encoding through the model.matrix function in R.

## RESULTS

### Demographic, health condition, and exposure characteristics

Patient-level summary statistics overall and by case-control status are summarized in Table 1. Of the 677 patients enrolled in the study, 222 (32%) had a positive *C. difficile* test during their hospitalization. Antibiotic use was common, with 67.7% of patients having received at least one antibiotic during their hospitalization. Eleven antibiotic classes were represented, with most patients having received cephalosporins (43.6%) followed by glycopeptide/lipopeptides (35.6%). Amount of time on antibiotics varied among patients, with a median length of 5 days [interquartile range (IQR)=13 days] and a median of 25% (IQR=66%) of a patient’s hospitalization spent on antibiotics. Distributions of the demographic characteristics were similar between cases and controls, while exposure variables and health conditions varied more substantially (Table 1).

**Table 1.**
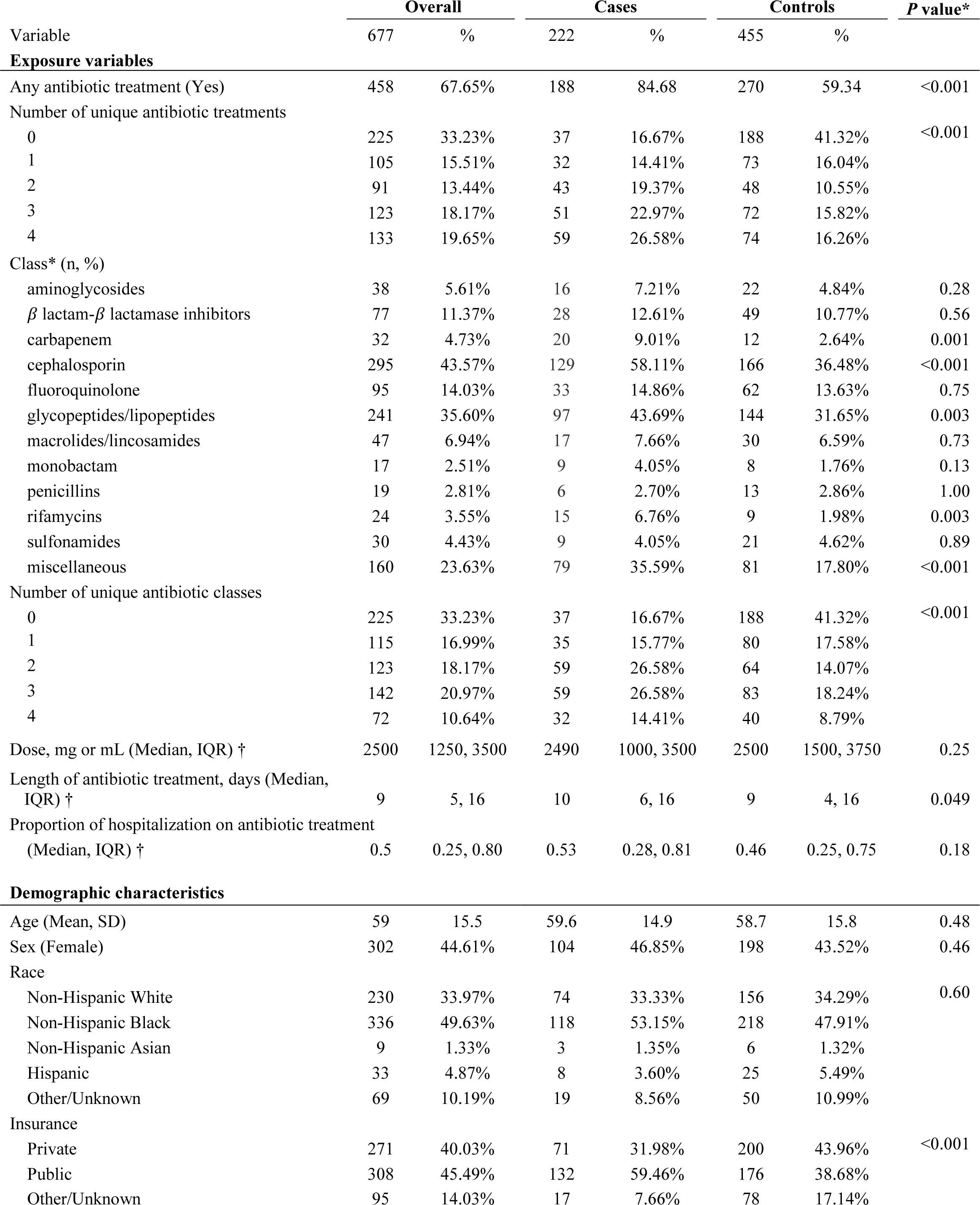
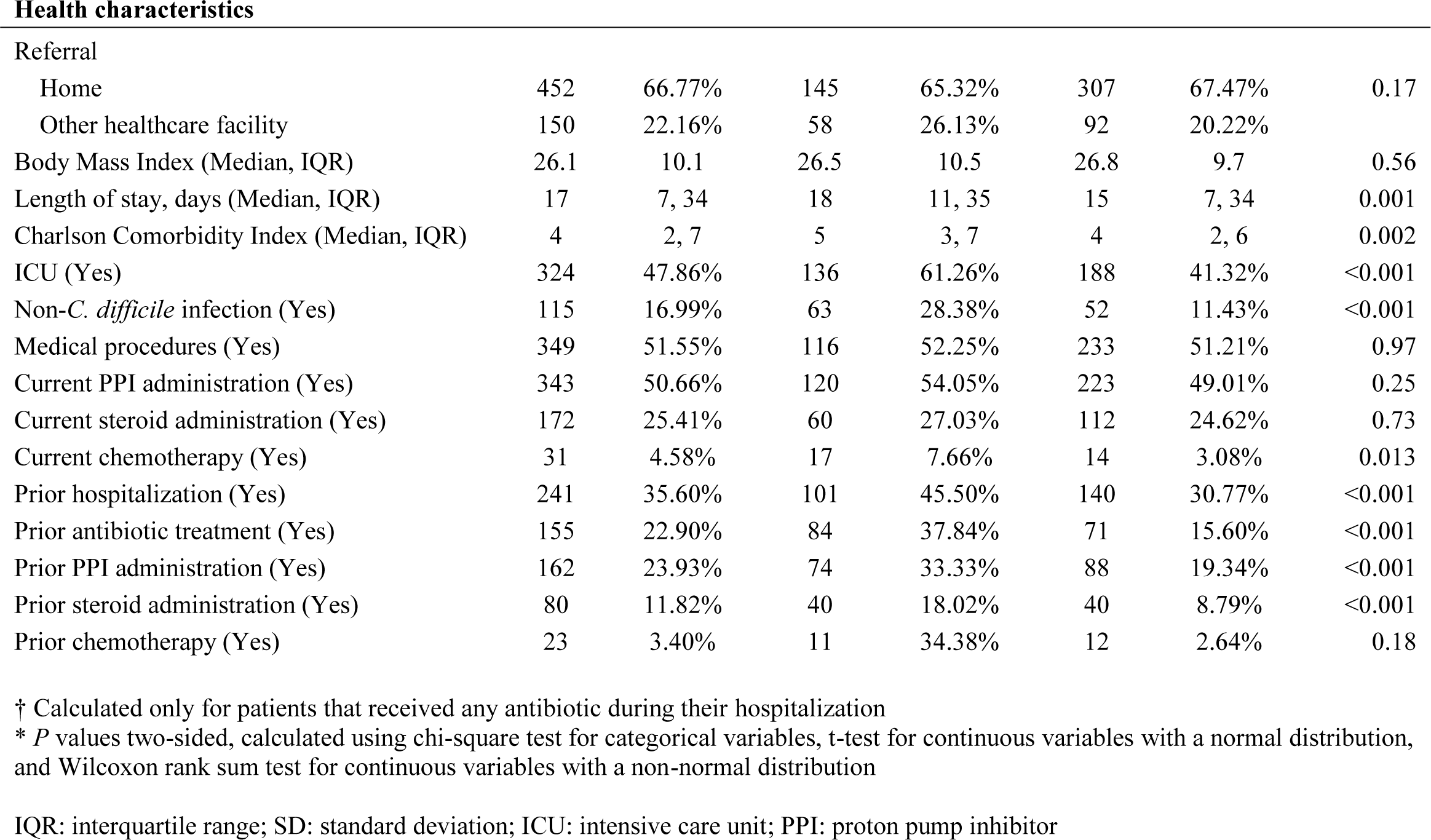
Distribution of patient-level univariate and bivariate characteristics overall and by case control status.

|  | Overall |  | Cases |  | Controls |  | P value* |
| --- | --- | --- | --- | --- | --- | --- | --- |
| Variable | 677 | % | 222 | % | 455 | % |  |
| Exposure variables |  |  |  |  |  |  |  |
| Any antibiotic treatment (Yes) | 458 | 67.65% | 188 | 84.68 | 270 | 59.34 | <0.001 |
| Number of unique antibiotic treatments |  |  |  |  |  |  |  |
| 0 | 225 | 33.23% | 37 | 16.67% | 188 | 41.32% | <0.001 |
| 1 | 105 | 15.51% | 32 | 14.41% | 73 | 16.04% |  |
| 2 | 91 | 13.44% | 43 | 19.37% | 48 | 10.55% |  |
| 3 | 123 | 18.17% | 51 | 22.97% | 72 | 15.82% |  |
| 4 | 133 | 19.65% | 59 | 26.58% | 74 | 16.26% |  |
| Class* (n, %) |  |  |  |  |  |  |  |
| aminoglycosides | 38 | 5.61% | 16 | 7.21% | 22 | 4.84% | 0.28 |
| $\beta$ lactam-β lactamase inhibitors | 77 | 11.37% | 28 | 12.61% | 49 | 10.77% | 0.56 |
| carbapenem | 32 | 4.73% | 20 | 9.01% | 12 | 2.64% | 0.001 |
| cephalosporin | 295 | 43.57% | 129 | 58.11% | 166 | 36.48% | <0.001 |
| fluoroquinolone | 95 | 14.03% | 33 | 14.86% | 62 | 13.63% | 0.75 |
| glycopeptides/lipopeptides | 241 | 35.60% | 97 | 43.69% | 144 | 31.65% | 0.003 |
| macrolides/lincosamides | 47 | 6.94% | 17 | 7.66% | 30 | 6.59% | 0.73 |
| monobactam | 17 | 2.51% | 9 | 4.05% | 8 | 1.76% | 0.13 |
| penicillins | 19 | 2.81% | 6 | 2.70% | 13 | 2.86% | 1.00 |
| rifamycins | 24 | 3.55% | 15 | 6.76% | 9 | 1.98% | 0.003 |
| sulfonamides | 30 | 4.43% | 9 | 4.05% | 21 | 4.62% | 0.89 |
| miscellaneous | 160 | 23.63% | 79 | 35.59% | 81 | 17.80% | <0.001 |
| Number of unique antibiotic classes |  |  |  |  |  |  |  |
| 0 | 225 | 33.23% | 37 | 16.67% | 188 | 41.32% | <0.001 |
| 1 | 115 | 16.99% | 35 | 15.77% | 80 | 17.58% |  |
| 2 | 123 | 18.17% | 59 | 26.58% | 64 | 14.07% |  |
| 3 | 142 | 20.97% | 59 | 26.58% | 83 | 18.24% |  |
| 4 | 72 | 10.64% | 32 | 14.41% | 40 | 8.79% |  |
| Dose, mg or mL (Median, IQR) † | 2500 | 1250, 3500 | 2490 | 1000, 3500 | 2500 | 1500, 3750 | 0.25 |
| Length of antibiotic treatment, days (Median, IQR) † | 9 | 5, 16 | 10 | 6, 16 | 9 | 4, 16 | 0.049 |
| Proportion of hospitalization on antibiotic treatment (Median, IQR) † | 0.5 | 0.25, 0.80 | 0.53 | 0.28, 0.81 | 0.46 | 0.25, 0.75 | 0.18 |
| Demographic characteristics |  |  |  |  |  |  |  |
| Age (Mean, SD) | 59 | 15.5 | 59.6 | 14.9 | 58.7 | 15.8 | 0.48 |
| Sex (Female) | 302 | 44.61% | 104 | 46.85% | 198 | 43.52% | 0.46 |
| Race |  |  |  |  |  |  |  |
| Non-Hispanic White | 230 | 33.97% | 74 | 33.33% | 156 | 34.29% | 0.60 |
| Non-Hispanic Black | 336 | 49.63% | 118 | 53.15% | 218 | 47.91% |  |
| Non-Hispanic Asian | 9 | 1.33% | 3 | 1.35% | 6 | 1.32% |  |
| Hispanic | 33 | 4.87% | 8 | 3.60% | 25 | 5.49% |  |
| Other/Unknown | 69 | 10.19% | 19 | 8.56% | 50 | 10.99% |  |
| Insurance |  |  |  |  |  |  |  |
| Private | 271 | 40.03% | 71 | 31.98% | 200 | 43.96% | <0.001 |
| Public | 308 | 45.49% | 132 | 59.46% | 176 | 38.68% |  |
| Other/Unknown | 95 | 14.03% | 17 | 7.66% | 78 | 17.14% |  |
| Health characteristics |  |  |  |  |  |  |  |
| Referral |  |  |  |  |  |  |  |
| Home | 452 | 66.77% | 145 | 65.32% | 307 | 67.47% | 0.17 |
| Other healthcare facility | 150 | 22.16% | 58 | 26.13% | 92 | 20.22% |  |
| Body Mass Index (Median, IQR) | 26.1 | 10.1 | 26.5 | 10.5 | 26.8 | 9.7 | 0.56 |
| Length of stay, days (Median, IQR) | 17 | 7, 34 | 18 | 11, 35 | 15 | 7, 34 | 0.001 |
| Charlson Comorbidity Index (Median, IQR) | 4 | 2, 7 | 5 | 3, 7 | 4 | 2, 6 | 0.002 |
| ICU (Yes) | 324 | 47.86% | 136 | 61.26% | 188 | 41.32% | <0.001 |
| Non-*C. difficile* infection (Yes) | 115 | 16.99% | 63 | 28.38% | 52 | 11.43% | <0.001 |
| Medical procedures (Yes) | 349 | 51.55% | 116 | 52.25% | 233 | 51.21% | 0.97 |
| Current PPI administration (Yes) | 343 | 50.66% | 120 | 54.05% | 223 | 49.01% | 0.25 |
| Current steroid administration (Yes) | 172 | 25.41% | 60 | 27.03% | 112 | 24.62% | 0.73 |
| Current chemotherapy (Yes) | 31 | 4.58% | 17 | 7.66% | 14 | 3.08% | 0.013 |
| Prior hospitalization (Yes) | 241 | 35.60% | 101 | 45.50% | 140 | 30.77% | <0.001 |
| Prior antibiotic treatment (Yes) | 155 | 22.90% | 84 | 37.84% | 71 | 15.60% | <0.001 |
| Prior PPI administration (Yes) | 162 | 23.93% | 74 | 33.33% | 88 | 19.34% | <0.001 |
| Prior steroid administration (Yes) | 80 | 11.82% | 40 | 18.02% | 40 | 8.79% | <0.001 |
| Prior chemotherapy (Yes) | 23 | 3.40% | 11 | 34.38% | 12 | 2.64% | 0.18 |

† Calculated only for patients that received any antibiotic during their hospitalization
\* *P* values two-sided, calculated using chi-square test for categorical variables, t-test for continuous variables with a normal distribution, and Wilcoxon rank sum test for continuous variables with a non-normal distribution
IQR: interquartile range; SD: standard deviation; ICU: intensive care unit; PPI: proton pump inhibitor

### Factor analysis of mixed data

Many of the antibiotic exposure measurements were correlated within our data (Figure 1). The variables with the most strongly correlated pairs included *number of unique treatments* with *number of unique classes* (corr=0.97), *any antibiotic treatment* (corr=0.78), and *dose* (corr=0.84); *number of unique classes* with *any antibiotic treatment* (corr=0.78) and *dose* (corr=0.81).

**Figure 1.**
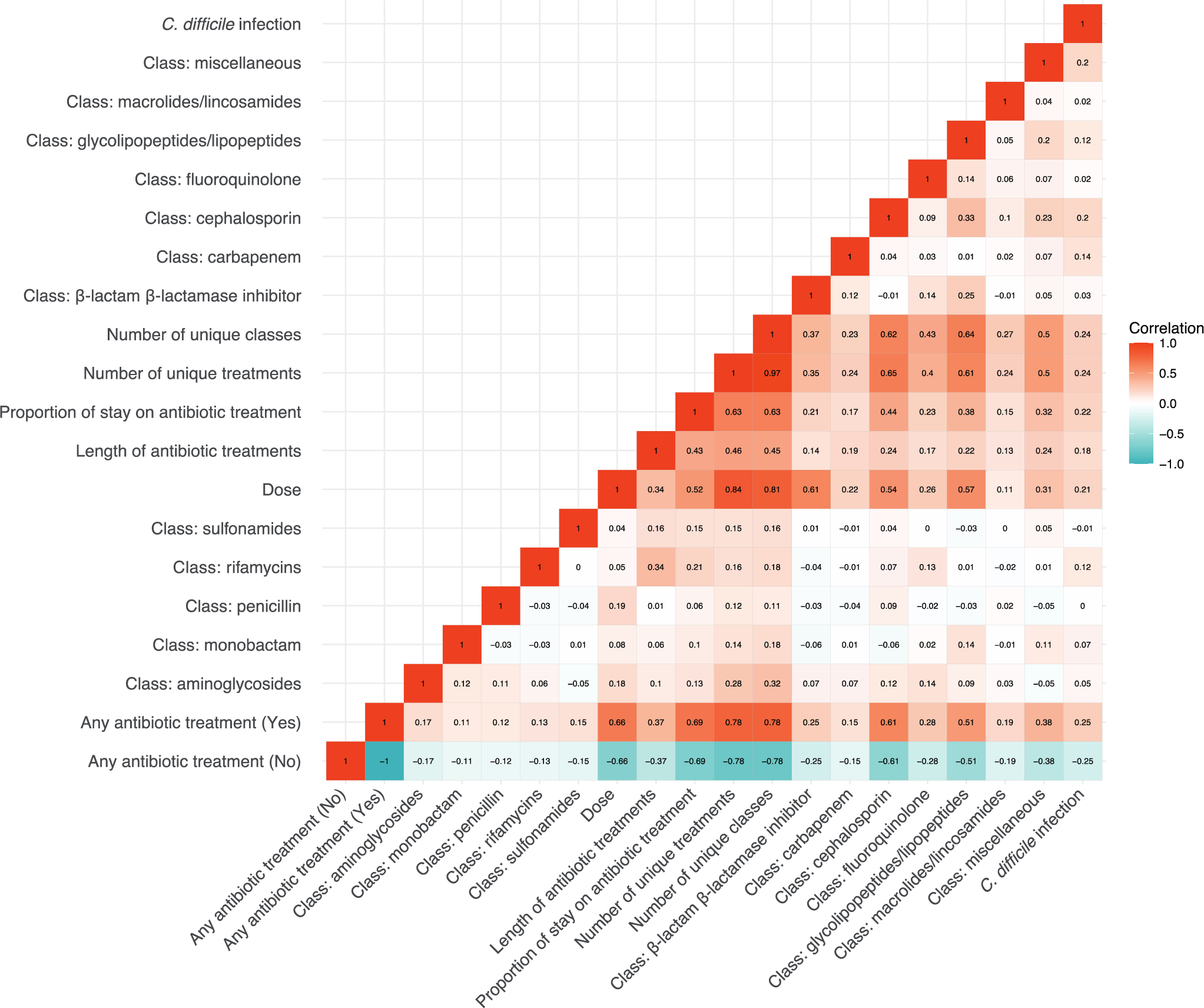
Correlation matrix among all antibiotic exposure variables and outcome variable (Clostridioides difficile infection) among a hospital-based case-control study population in Philadelphia, Pennsylvania from 2014-2018. Categorical variables were transformed to numeric using one-hot encoding.

The factor analysis identified six factors (dimensions) with eigenvalues ≥1, considering the correlations and variation within our dataset among the antibiotic exposure variables alone. Considering all six dimensions, the variables that contribute the most to each dimension are illustrated in Figure 2, which plots the total contribution of the top variables. The red line represents the expected average contribution if all contributions were uniform across the variables.

**Figure 2.**
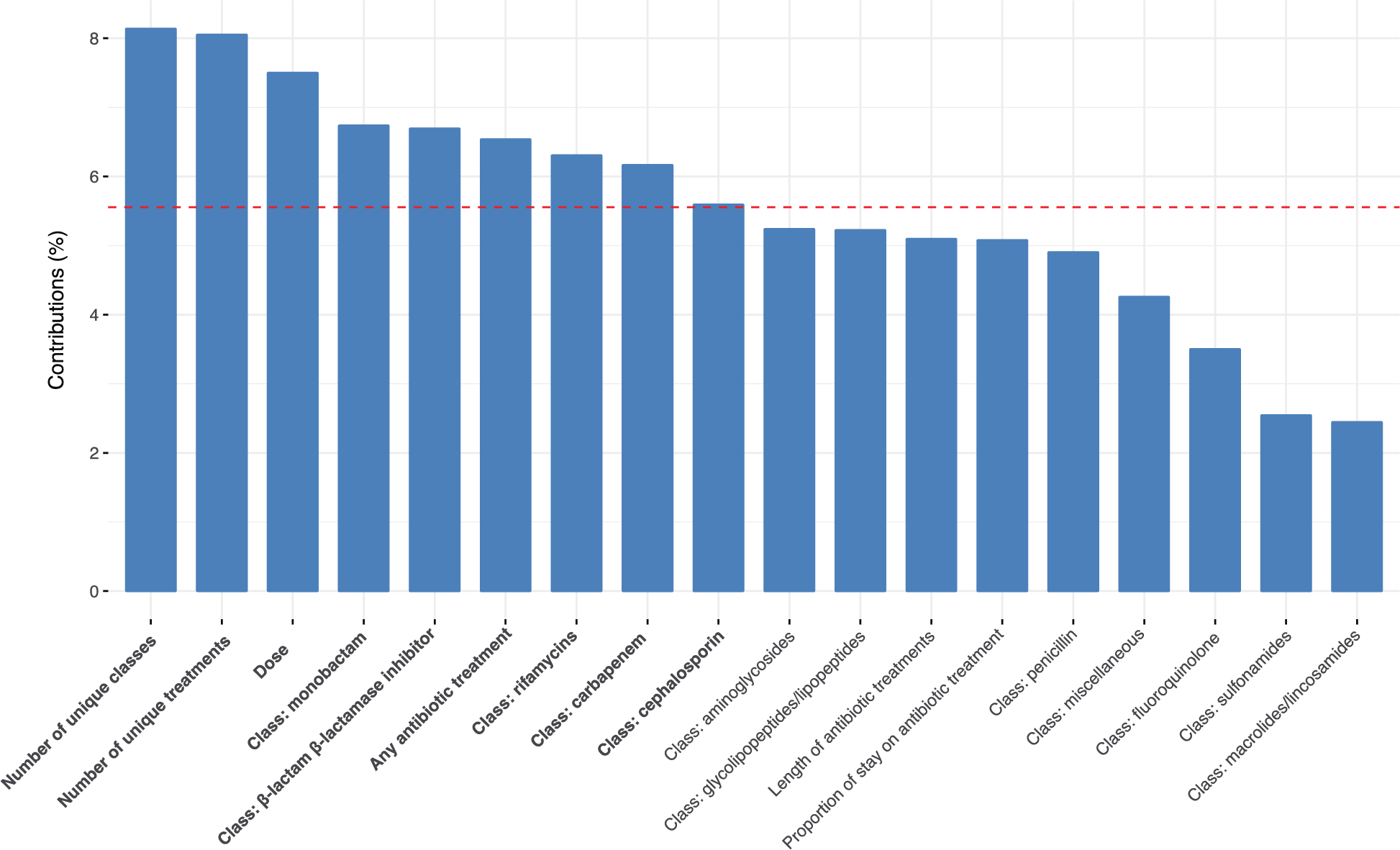
Plot of the contributions of antibiotic exposure variables to the top six dimensions created from a factor analysis of mixed data within a hospital-based case-control study population in Philadelphia, Pennsylvania from 2014-2018. The red line represents the expected average contribution if all contributions were uniform across the variables.

The variables with the greatest contributions to the six dimensions include 1) *number of unique classes*, 2) *number of unique treatments*, 3) *dose*, 4) monobactam class, 5) β lactam-β lactamase inhibitors class, 6) *any antibiotic treatment*, 7) rifamycin class, 8) carbapenem class, and 9) cephalosporin class.

### Logistic regression models

Results from the seven logistic regression models can be found in Table 2. All regression models were adjusted for the following variables, according to the results of the bivariate analyses and using a *P* value cut-off of 0.1: insurance, length of stay, Charlson Comorbidity Index, ICU, non-*C. difficile* infection, current chemotherapy, prior hospitalization, prior antibiotics, prior PPI exposure, and prior steroid exposure. The two interaction terms were also included in all models. The model with the lowest AIC used the *any antibiotic treatment* variable to represent antibiotic exposure (AIC=719.7), followed by the model that used the *proportion of hospitalization on antibiotic treatment* variable (AIC=720.8). The remaining models had similar AIC values (AIC range=725-728). The odds of testing positive for *C. difficile* infection were increased for patients that experienced these two exposure measures [Odds Ratio (OR)_any treatment_ = 2.02, 95% Confidence Interval (CI)=1.27, 3.24; OR_proportion of hospitalization_ = 2.19, 95% CI=1.27, 3.78] compared to patients that did not experience these exposure measures, respectively, adjusting for the previously described potential confounders.

**Table 2.** Logistic regression modeling of odds of *Clostridioides difficile* infection by seven measurements of antibiotic exposure.

| Variable | OR | 95% CI |  | P value* | AIC |
| --- | --- | --- | --- | --- | --- |
|  |  | LL | UL |  |  |
| Exposure variables |  |  |  |  |  |
| Any antibiotic treatment | 2.02 | 1.27 | 3.24 | 0.003 | 719.67 |
| Number of unique antibiotic treatments | 1.14 | 0.99 | 1.30 | 0.071 | 725.51 |
| Class $\Delta$ | | | | | 726.90 |
| aminoglycosides | 0.85 | 0.36 | 1.92 | 0.70 |  |
| $\beta$ lactam-β lactamase inhibitors | 0.70 | 0.37 | 1.32 | 0.28 | |
| carbapenem | 2.16 | 0.90 | 5.30 | 0.086 |  |
| cephalosporin | 1.61 | 1.06 | 2.44 | 0.024 |  |
| fluoroquinolone | 0.77 | 0.44 | 1.31 | 0.33 |  |
| glycopeptides/lipopeptides | 0.88 | 0.57 | 1.37 | 0.58 |  |
| macrolides/lincosamides | 0.96 | 0.47 | 1.92 | 0.91 |  |
| monobactam | 2.07 | 0.61 | 7.26 | 0.25 |  |
| penicillins | 0.80 | 0.25 | 2.33 | 0.69 |  |
| rifamycins | 2.12 | 0.79 | 5.88 | 0.14 |  |
| sulfonamides | 0.61 | 0.24 | 1.45 | 0.28 |  |
| miscellaneous | 1.78 | 1.15 | 2.76 | 0.009 |  |
| Number of unique antibiotic classes | 1.14 | 0.98 | 1.32 | 0.096 | 726.00 |
| Dose (grams or liters) | 1.06 | 0.95 | 1.18 | 0.27 | 727.57 |
| Length of antibiotic treatment, days | 1.01 | 1.00 | 1.03 | 0.13 | 726.16 |
| Proportion of hospitalization on antibiotic treatment | 2.19 | 1.27 | 3.78 | 0.005 | 720.78 |

All models were adjusted for insurance, length of stay, Charlson Comorbidity Index, intensive care unit, non-*C. difficile* infection, current chemotherapy, prior hospitalization, prior antibiotic treatment, prior proton pump inhibitor administration, prior steroid administration, and interaction terms between current and prior antibiotics and PPI.
OR: odds ratio; CI: confidence interval; LL: lower limit; UL: upper limit; AIC: Akaike Information Criterion
\* Two-sided P values
$\Delta$ class models were adjusted for all other classes
§ proportion of days on antibiotic treatment was not adjusted for length of stay (used in variable calculation)

### LASSO regression

The primary LASSO regression analysis (λ min) identified 22 covariates to be included in the predictive model for *C. difficile* infection: exposure variables *any antibiotic treatment*, class variables β lactam-β lactamase inhibitors, carbapenem, cephalosporin, fluoroquinolone, monobactam, rifamycin, sulfonamides, and miscellaneous, *proportion of hospitalization on antibiotic treatment*; and covariates sex, race, insurance, ICU, non-*C. difficile* infection, medical procedures, current steroid exposure, current chemotherapy, prior antibiotics, prior PPI exposure, prior steroid exposure, and the interaction between current antibiotics × current PPI. When running the LASSO regression model using the alternative λ value, 10 variables were selected to be retained in the model. The primary LASSO regression model (λ min) had superior model fit (AIC=519.6 vs. 587.5) and predictive performance (AUC=0.78 vs. 0.75; sensitivity=0.41 vs. 0.25; and accuracy=0.69 vs. 0.66). The coefficients, fit, and performance of each model are detailed in Table 3.

**Table 3.**
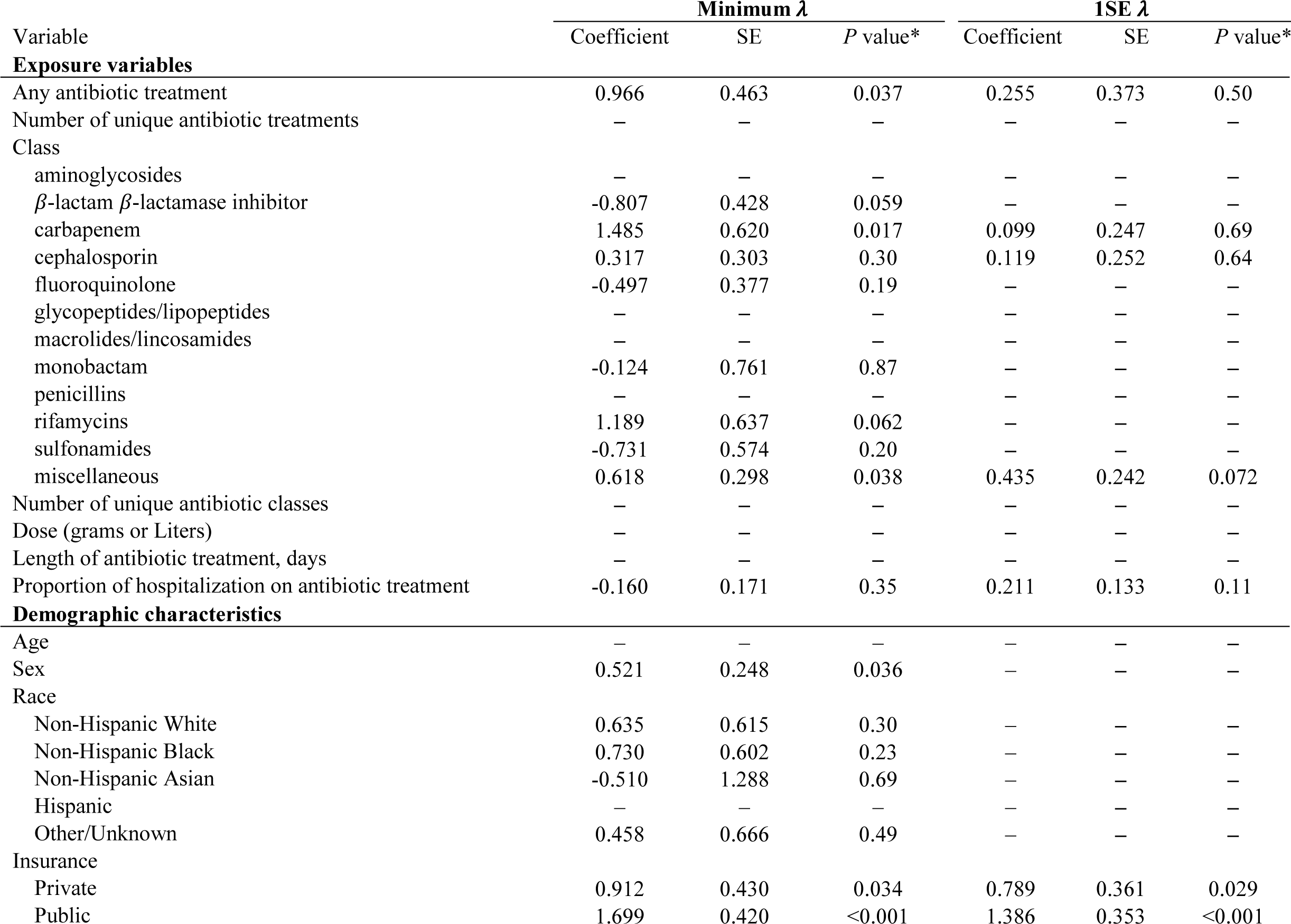
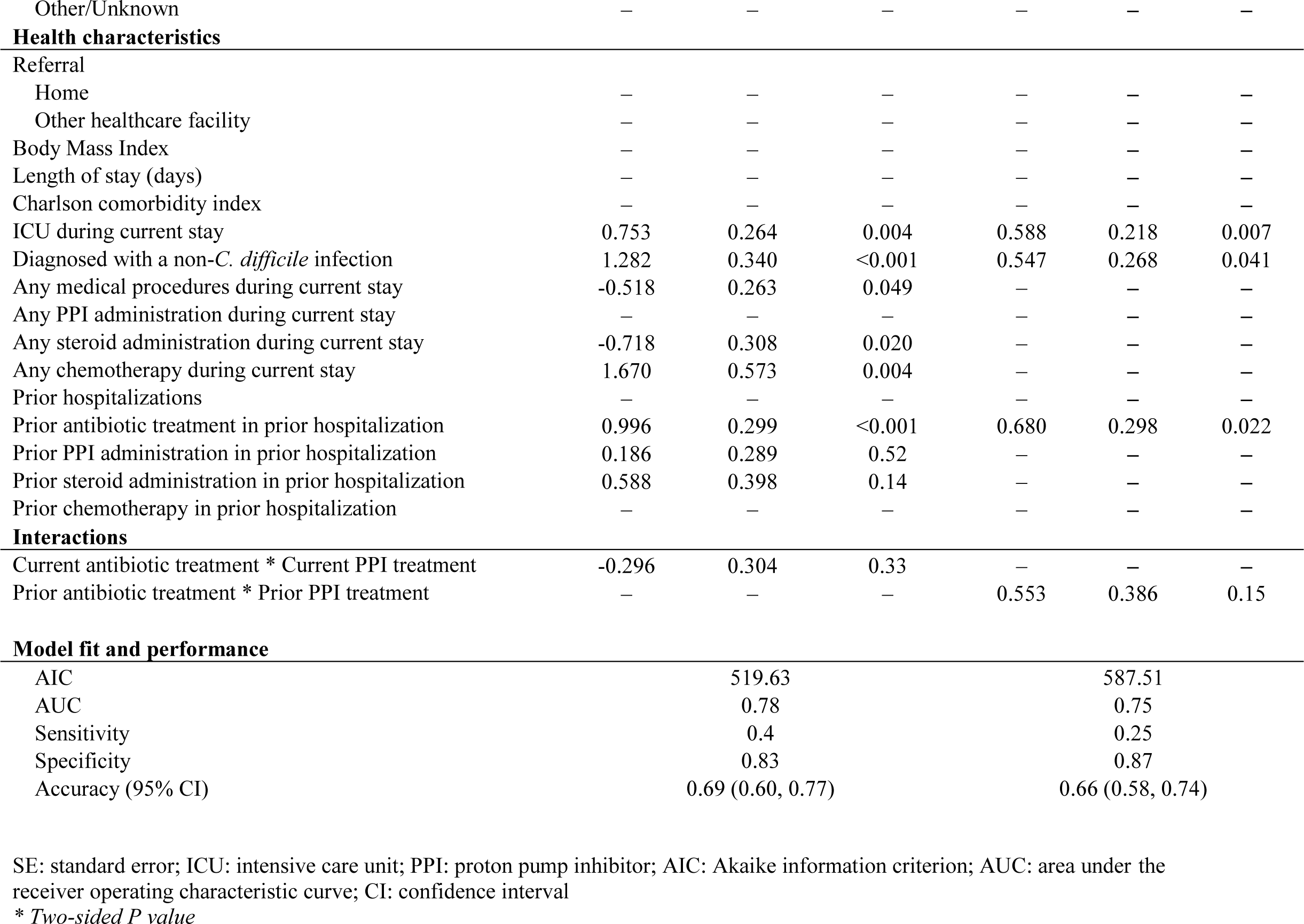
Regression coefficients, fit, and performance of predictive models for hospital-associated *Clostridioides difficile* infection using variables selected by LASSO regression using 1) the minimum value of λ, and 2) the maximum value of λ within 1 standard error of the minimum λ.

## DISCUSSION

This research sought to summarize the most common ways antibiotic exposure is measured and operationalized in clinical and epidemiologic research and provide examples of analytic methods as a resource for investigators when determining how to best capture this exposure within the context of their research. We used three approaches to explore and identify antibiotic exposure variables of importance to our data (factor analytics), and in predicting hospital-associated *C. difficile* infection (logistic and LASSO regression analytics). The factor analysis identified 9 variables contributing the most variation in the dataset: *any antibiotic treatment*; *number of classes*; *number of treatments*; *dose*; and class variables *monobactam*, β *lactam-*β *lactamase inhibitors*, *rifamycin*, *carbapenem*, and *cephalosporin*. The logistic regression models resulting in the best model fit used predictors *any antibiotic exposure* and *proportion of hospitalization on antibiotics*. The LASSO model selected 22 variables for inclusion in the predictive model, with exposure variables including *any antibiotic treatment*; class variables β *lactam-*β *lactamase inhibitors*, *carbapenem*, *cephalosporin*, *fluoroquinolone*, *monobactam*, *rifamycin*, *sulfonamides*, and *miscellaneous*; and *proportion of hospitalization on antibiotic treatment*.

In all three of our analytic approaches, the variable representing *any antibiotic exposure* was selected as an appropriate analytic choice – in the factor analysis this variable was among the top contributing variables to the selected factors, in the logistic regression method the model using this exposure variable was the best fit to the data, and in the LASSO regression method this variable was selected to remain in the model under both penalty scenarios. Logically, these results are driven by the data and the outcome of interest. And while we provide some examples of analytic methods for variable selection, they should be used in conjunction with or as a complement to a theoretical approach to variable selection. Nonetheless, we believe that it is beneficial to recognize that each of these aspects of antibiotic treatment provides a different measure of exposure, which has the potential to impact the final effect estimates in an associational or causal analysis.

As with any analytic model, there will always be a tradeoff when it comes to balancing bias and variance. Although the *any antibiotic treatment* variable performed well in all approaches, using only one measure may result in loss of information in another aspect. For example, using a dichotomous response for *any antibiotic treatment* over the course of the hospitalization will miss information relating to amount of exposure, with which medication, and the method of exposure. We recognize that some investigators may not have access to underlying antibiotic data, and certain sources even when accessible, may be more reliable than others. For example, while bar code administration is often considered the gold standard in antibiotic use measurement,^29,30^ it may not be as easy to collect if there are different means for documentation or the EMR is not directly accessible to the researcher. Thus measurement error remains a distinct possibility and we refer readers to quantitative bias analysis techniques to understand the impact of this information bias.^31^

To combat both varying results across different measures and potential for missing information if using only one measure, much of the research that we reviewed conducted multiple analyses, often providing conflicting results. No clear gold standard for measuring antibiotic exposure currently exists, and while multiple models can provide a range of individual effect estimates (hopefully encompassing the true effect estimate), this does not necessarily explain which measure is the most appropriate fit, or how the collective aspects of antibiotics can affect an outcome. This paper provides guidance for researchers working with complex exposures such as antibiotic treatment, with potential solutions for identifying an ideal exposure characterization depending on data availability, access to certain modeling tools, and research objectives. Another option, which could be considered an extension of the factor analysis approach, could be to develop a composite or calculated variable, which combine information based on multiple aspects of treatment into one single variable.^9,21,32,33^ Future research should work to develop similar measures that can account for multiple aspects across fields of clinical research and epidemiology.

### Limitations

These methods and analytic approaches are not without limitations. First, the data that are considered in this research are limited to summary measures of exposure, or time-invariant measures. Considering time-varying exposure may be important in predicting certain outcomes, and if a patient is on multiple treatments, it may be preferable to use longitudinal methods to capture changes in exposure over time. Another potential limitation to this research is that we considered only one method of variable selection for the final predictive model. LASSO is a regression-based machine learning method, selected based on the intuitiveness of its methods and results, and the model’s ability to perform well with smaller datasets.^34,35^ Other regression-based and tree-based machine learning methods exist,^36–39^ in addition to non-machine learning variable selection methods.^40^ It is important to understand the differences in the methods and assumptions for each variable selection model when choosing an analytic approach.^37^

Although the objective of this paper was to provide a demonstration of analytic approaches for variable exploration and selection, and not necessarily to report on the results of the analyses, we nonetheless believe it is important to mention the potential lack of generalizability of our results to other health systems due to the unique patient population served by this hospital. Vader et al was the first to describe results from this study, and identified differences in race/ethnicity and age of cases in our population compared to nationally representative samples. ^23^ We advise readers to interpret the results with these considerations in mind.

## Conclusion

Investigators studying antibiotic exposure should consider multiple aspects of treatment informed by their research question and the theory on how antibiotics may impact the distribution of the outcome in their target population. Failure to consider the complexities of antibiotic exposure measurement may lead to exposure misclassification, potentially biasing associations between antibiotic use and the outcome of interest. Improved understanding of the specific aspects of antibiotic exposure that increases a patient’s risk of secondary complications or diseases will lead to improved patient outcomes and more informative preventive measures.

## Data Availability

Data not available - participant consent.

https://github.com/jlywebster/abxexp

**Supplement 1:** Methods and results of a methodological rapid review on commonly used measurements of antibiotic exposure in clinical research.

## Methods

PubMed was searched for articles published in English within the last 20 years (2003-2023) that included both “antibiotic” and either “exposure” or “use” in the title and included the keywords “patient” or “hospital” to eliminate non-human studies. Studies that were not observational or clinical, that did not include human subjects, or that evaluated antibiotic therapy as an outcome were excluded from the search.

## Results

### Methods used to measure, operationalize, and analyze antibiotic exposure

The methodological rapid review identified 92 papers, of which 81 (88%) focused on patient-level outcomes and 11 (12%) focused on hospital-level outcomes (citations of the papers selected for full review are available in Appendix 1). The review of the literature found that a major driver in the decision for how investigators operationalized antibiotic use depended on the outcome of interest. Broadly speaking, these outcomes could be categorized as 1) individual or patient-level outcomes, and 2) aggregate or hospital-level outcomes. Patient-level outcomes could further be subcategorized into infectious outcomes (including *C. difficile* infection, sepsis, infections resistant to antibiotics) or immune-mediated disorders (including asthma, arthritis, psoriasis) – outcomes within these two categories have been found to be positively associated with antibiotic use. Other patient-level outcomes that did not fit into either subcategory included mental health outcomes, weight-related outcomes, and general microbiome/antimicrobial resistant outcomes. Regardless of the outcome of interest, the most commonly cited theory relating patient-level antibiotic exposure to disease involves the disruption of the human microbial communities.[1] Hospital-level outcomes focused on antibiotic stewardship and an aggregate of related patient-level outcomes. Studies that focused on antimicrobial resistance and the general effect of antibiotic exposure on the human microbiome used both individual and hospital-level data.

### Patient-level outcomes

Of the 92 papers reviewed, 42 (46%) focused on individual, infectious disease-related outcomes. Literature that focused on the relationship between antibiotic exposure and patient-level infections used a variety of methods to measure and operationalize antibiotic exposure. Most of these studies (28, 66%) acknowledged the potential for differences in effect measures based on definition of antibiotic exposure, and thus employed multiple methods within a single study.[2–5] Any exposure (yes/no) to any antibiotic or any exposure (yes/no) to specific medications were used the most frequently (19, 45%). Specific medications of interest necessarily varied based on the pathogen of interest (*C. difficile*, *E. coli*, etc.) and many of these studies grouped the specific medication by class (16, 38%) or spectrum (1, 2%). The choice of specific medications and categorizations were chosen by the investigators based on previous literature and expert knowledge, although this was not always explicitly stated. Other frequently used methods were timing of treatment (within hospital stay, relative to disease onset, etc.) (17, 41%), [6–8] and duration of treatment (13, 31%), often described as “days of therapy” (DOT), calculated as the cumulative number of days a patient was exposed to any antibiotics over the course of follow-up.[9–12] Less common approaches included risk level of antibiotic (specific to outcome of interest) (4, 10%), [3, 4, 13, 14] number of unique antibiotic courses (3, 7%), [4, 5, 14] and dose (3, 7%).[4, 5, 15]

The review identified 26 (28%) studies that focused on immune-mediated disorders as their outcome of interest. Similar to studies focusing on infections, those that assessed the association between antibiotic exposure and immune-mediated disorders considered the role that antibiotics play in the disruption of the human microbiome. These studies tended to focus on childhood exposures, and often during a time-period specific to a child’s development (i.e., first two years of a child’s life, two years prior to diagnosis of the outcome of interest) (20, 77%).[16–18] As such, much of the literature in this domain (17, 65%) operationalized antibiotic exposure as any exposure (yes/no) or any exposure (yes/no) to specific medications within specified windows of time.[19–22] 6 (23%) studies quantified how many exposures (unique treatment courses) occurred total or within specified time periods.[20, 23–25] Class of medication was also used as an exposure measurement in 11 (42%) studies, with groupings based on type of antibiotic [anti-anaerobic vs. non-anti-anaerobic (1, 4%) or spectrum (1, 4%)].[24, 26]

### Hospital-level outcomes

Our review identified 11 (12%) studies that focused on aggregated measures of antibiotic exposure. Investigators that conduct research on antibiotic use as a predictor of antibiotic resistance, or in conjunction with a hospital’s antimicrobial stewardship program, typically utilized hospital-level data that measured antibiotic use over a certain period of time. Validated measures have been developed to systematically measure antibiotic prescriptions in hospitals, in an effort to identify unnecessary antibiotic use and improve patient outcomes. The most common measure identified in the literature was the defined daily dose (DDD), developed by the World Health Organization, defined as “the assumed average maintenance dose per day for a drug used for its main indication in adults.”[27] Studies using this measure will typically take the number of grams of each antibiotic used (purchased, dispensed, administrated) summed over a certain period of time, and divided by the DDD. The DDD is ideal for studies that evaluate antibiotic use on an aggregate hospital level and utilize antibiotics that have been assigned an Anatomical Therapeutic Chemical (ATC) code (DDDs have only been calculated for medicines given an ATC code). The majority (8, 72%) of studies that we reviewed used some form of the DDD measure to quantify antibiotic use.[28–30] Aggregated duration of therapy was used in 6 (55%) studies to measure the average number of days patients were exposed to antibiotics.[31–33] Duration of therapy was typically measuring using DOT – a measure that is increasingly recommended (and sometimes required) over DDD when measuring antibiotic use for stewardship efforts.[34] Leung and colleagues [35] published a comprehensive systematic review detailing additional methods that have been used to measure antibiotic exposure on a hospital level.

---

**Supplemental Table 1.** Description of important characteristics of antibiotic exposure, including suggested variable type (operationalization), data requirements for measurement, advantages, and disadvantages.

| *Characteristic* | *Description* | *Variable type* | *Data requirements* | *Advantages & Disadvantages* |
| --- | --- | --- | --- | --- |
| *Measurements* |  |  |  |  |
| *Receipt of any antibiotic* | Whether or not the patient received an antibiotic treatment. | Dichotomous (Yes / No) | Dependent on level of measurement for time: patient-level data on *any* antibiotic treatment over entire hospitalization; day-level data on antibiotic treatment for day-specific (longitudinal) exposure measurement. | + Feasibility, simple to collect and analyze. Smallest unit of measurement (if collected at the day-level), ability to create summary measures based on data.<br>– Requires calculation of summary measures to determine amount of exposure (length of time, number of courses). Unable to determine dosage, type of exposure (medication), or method of exposure. Daily (longitudinal) data may be difficult to collect and analyze. |
| *Medication* | The specific antibiotic medication that the patient received during their hospitalization. Sub-categories: class, generation, spectrum. | Categorical | Builds off *Receipt of any antibiotic*. Additional requirement includes name of the specific medication administered for each measure of antibiotic receipt. Could be collapsed or further classified into class, spectrum, generation. | + Important detail regarding type of antibiotic exposure.<br>– Unable to determine amount of exposure or method of exposure, potential difficulty in determining meaningful categorizations or classifications, small cell sizes if many medications are prescribed. |
| *Dosage* | The dosage of antibiotic treatment that the patient received over the course of their hospitalization. | Continuous, categorical | Builds off *Receipt of any antibiotic*. Additional requirement includes the specific dosage administered for each measure of antibiotic receipt. | + important detail regarding amount of exposure<br>– Only one aspect of amount of exposure, unable to determine length of time on antibiotics or whether different courses were administered, type of exposure, or method of exposure. |
| *Route of administration* | The route of administration for the antibiotic treatment that the patient received over the course of their hospitalization. | Categorical | Builds off *Receipt of any antibiotic*. Additional requirement includes the specific route of administration for each measurement of antibiotic receipt. | + Important detail regarding method of exposure.<br>– Unable to determine amount or type of exposure. |
| *Summaries* |  |  |  |  |
| *Number of antibiotic courses* | Cumulative number (sum) of unique antibiotic courses a patient received over the course of their hospitalization. | Continuous, categorical | Summary measure of *Receipt of any antibiotic*. Day-level data required to distinguish between unique antibiotic courses. Unique courses can be determined by *Medication* and timing of treatment. | + Ability to determine amount of exposure to an extent. Summary measure is useful for comparing overall exposure patterns.<br>– Only one aspect of amount of exposure, unable to determine length of time or dosage, type of exposure, or method of exposure. |
| *Length of time on antibiotics* | Cumulative amount (sum) of time (days, hours) that a patient received antibiotics over the course of their hospitalization. Alternative forms could include average length of time per unique antibiotic course, proportion of hospitalization on antibiotic treatment. | Continuous, proportion | Summary measure of *Receipt of any antibiotic*. Day-level data required to calculate the number of days the patient received antibiotics. This measure can be calculated for all courses (total number of days on antibiotic treatment), per unique course (average length of antibiotic course), as a proportion of total length of stay (total number of days on antibiotic treatment divided by length of stay). | + Ability to determine amount of exposure to an extent. Summary measure is useful for comparing overall exposure patterns.<br>– Only one aspect of amount of exposure, unable to determine the dosage, type of exposure, or method of exposure. |

**Supplemental Table 2.** Antibiotic class categorizations and the specific medication(s) that comprise them.

| **Class** | **Specific medication(s)** |
| --- | --- |
| **aminoglycosides** | amikacin, gentamicin, tobramycin, neomycin |
| **β-lactam β-lactamase inhibitors** | ampicillin-sulbactam, amoxicillin-clavulanate, piperacillin-tazobactam, ticarcillin-clavulanate |
| **carbapenem** | ertapenem, imipenem, meropenem |
| **cephalosporin** | cefazolin, cefepime, ceftriaxone, ceftaroline, ceftazidime-avibactam, ceftazidime, ceftolozane-tazobactam, cefuroxime, cephalexin, ceftazidime-cefepime |
| **fluoroquinolone** | ciprofloxacin, levofloxacin, moxifloxacin |
| **glycolipopeptides/lipopeptides** | vancomycin, daptomycin |
| **macrolides/lincosamides** | azithromycin, erythromycin, clarithromycin, clindamycin |
| **monobactam** | aztreonam |
| **penicillin** | ampicillin, amoxicillin, nafcillin, oxacillin, penicillin g |
| **tetracyclines** | tetracycline, doxycycline, tigecycline, minocycline |
| **rifamycins** | rifaximin, rifampin |
| **sulfonamides** | trimethoprim-sulfamethoxazole |
| **miscellaneous** | dapsone, ethambutol, metronidazole, colistimethate, isoniazid, linezolid, nitrofurantoin, mitomycin |

## References

1. Song J, Cohen B, Liu J, Larson E, Zachariah P. The Association Between the Frequency of Interruptions in Antibiotic Exposure and the Risk of Health Care-Associated Clostridiodes difficile Infection. Current Therapeutic Research. 2020/01/01/ 2020;93:100600. 10.1016/j.curtheres.2020.100600

2. Ben-Ami R, Olshtain-Pops K, Krieger M, et al. Antibiotic exposure as a risk factor for fluconazole-resistant Candida bloodstream infection. Antimicrobial agents and chemotherapy. 2012;56(5):2518–2523.

3. DiDiodato G, Fruchter L. Antibiotic exposure and risk of community-associated Clostridium difficile infection: A self-controlled case series analysis. Am J Infect Control. Jan 2019;47(1):9–12. doi:10.1016/j.ajic.2018.06.016

4. Hung YP, Lin HJ, Wu TC, et al. Risk factors of fecal toxigenic or non-toxigenic Clostridium difficile colonization: impact of Toll-like receptor polymorphisms and prior antibiotic exposure. PLoS One. 2013;8(7):e69577. doi:10.1371/journal.pone.0069577

5. Smith JT, Manickam RN, Barreda F, et al. Quantifying the breadth of antibiotic exposure in sepsis and suspected infection using spectrum scores. Medicine (Baltimore). Oct 14 2022;101(41):e30245. doi:10.1097/md.0000000000030245

6. Song J, Cohen B, Zachariah P, Liu J, Larson EL. Temporal change of risk factors in hospital-acquired Clostridioides difficile infection using time-trend analysis. Infection Control & Hospital Epidemiology. 2020;41(9):1048–1057. doi:10.1017/ice.2020.206

7. Stevens V, Dumyati G, Brown J, Wijngaarden E. Differential risk of Clostridium difficile infection with proton pump inhibitor use by level of antibiotic exposure. Pharmacoepidemiol Drug Saf. Oct 2011;20(10):1035–42. doi:10.1002/pds.2198

8. Zhao J, Murray S, LiPuma JJ. Modeling the Impact of Antibiotic Exposure on Human Microbiota. Scientific Reports. 2014/03/11 2014;4(1):4345. doi:10.1038/srep04345

9. Kuster SP, Rudnick W, Shigayeva A, et al. Previous antibiotic exposure and antimicrobial resistance in invasive pneumococcal disease: results from prospective surveillance. Clin Infect Dis. Oct 2014;59(7):944–52. doi:10.1093/cid/ciu497

10. Chapman TJ, Pham M, Bajorski P, Pichichero ME. Antibiotic Use and Vaccine Antibody Levels. Pediatrics. May 1 2022;149(5)doi:10.1542/peds.2021-052061

11. Horton DB, Scott FI, Haynes K, et al. Antibiotic exposure and juvenile idiopathic arthritis: a case–control study. Pediatrics. 2015;136(2):e333–e343.

12. Hoskin-Parr L, Teyhan A, Blocker A, Henderson A. Antibiotic exposure in the first two years of life and development of asthma and other allergic diseases by 7.5 yr: a dose-dependent relationship. Pediatric Allergy and Immunology. 2013;24(8):762–771.

13. Uldbjerg CS, Miller JE, Burgner D, Pedersen LH, Bech BH. Antibiotic exposure during pregnancy and childhood asthma: a national birth cohort study investigating timing of exposure and mode of delivery. Arch Dis Child. Sep 2021;106(9):888–894. doi:10.1136/archdischild-2020-319659

14. Yang SI, Lee E, Jung YH, et al. Effect of antibiotic use and mold exposure in infancy on allergic rhinitis in susceptible adolescents. Ann Allergy Asthma Immunol. Aug 2014;113(2):160–165.e1. doi:10.1016/j.anai.2014.05.019

15. Aris IM, Lin PD, Rifas-Shiman SL, et al. Association of Early Antibiotic Exposure With Childhood Body Mass Index Trajectory Milestones. JAMA Netw Open. Jul 1 2021;4(7):e2116581. doi:10.1001/jamanetworkopen.2021.16581

16. Block JP, Bailey LC, Gillman MW, et al. Early Antibiotic Exposure and Weight Outcomes in Young Children. Pediatrics. Dec 2018;142(6)doi:10.1542/peds.2018-0290

17. Prichett LM, Yolken RH, Wu L, Severance EG, Kumra T. Relationship between antibiotic exposure and subsequent mental health disorders in a primary care health system. Brain Behav Immun Health. May 2022;21:100430. doi:10.1016/j.bbih.2022.100430

18. Polk RE, Fox C, Mahoney A, Letcavage J, MacDougall C. Measurement of adult antibacterial drug use in 130 US hospitals: comparison of defined daily dose and days of therapy. Clinical infectious diseases. 2007;44(5):664–670.

19. Stanić Benić M, Milanič R, Monnier AA, et al. Metrics for quantifying antibiotic use in the hospital setting: results from a systematic review and international multidisciplinary consensus procedure. Journal of Antimicrobial Chemotherapy. 2018;73(suppl_6):vi50–vi58. doi:10.1093/jac/dky118

20. U.S. Department of Health and Human Services. 2021 National Healthcare Safety Network Antimicrobial Use Option Report. 2022. https://www.cdc.gov/nhsn/pdfs/datastat/2021-AU-Report-508.pdf

21. Gerber JS, Hersh AL, Kronman MP, Newland JG, Ross RK, Metjian TA. Development and Application of an Antibiotic Spectrum Index for Benchmarking Antibiotic Selection Patterns Across Hospitals. Infection Control & Hospital Epidemiology. 2017;38(8):993–997. doi:10.1017/ice.2017.94

22. McCarthy KN, Hawke A, Dempsey EM. Antimicrobial stewardship in the neonatal unit reduces antibiotic exposure. Acta Paediatr. Oct 2018;107(10):1716–1721. doi:10.1111/apa.14337

23. Vader DT, Weldie C, Welles SL, Kutzler MA, Goldstein ND. Hospital-acquired Clostridioides difficile infection among patients at an urban safety-net hospital in Philadelphia: Demographics, neighborhood deprivation, and the transferability of national statistics. Infect Control Hosp Epidemiol. Aug 2021;42(8):948–954. doi:10.1017/ice.2020.1324

24. Stevens V, Dumyati G, Brown J, van Wijngaarden E. Differential risk of Clostridium difficile infection with proton pump inhibitor use by level of antibiotic exposure. Pharmacoepidemiology and drug safety. 2011;20(10):1035–1042.

25. Ratner B. The correlation coefficient: Its values range between +1/−1, or do they? *Journal of Targeting*, Measurement and Analysis for Marketing. 2009/06/01 2009;17(2):139–142. doi:10.1057/jt.2009.5

26. Braeken J, van Assen MALM. An empirical Kaiser criterion. Psychological Methods. 2017;22(3):450–466. doi:10.1037/met0000074

27. Abdi H, Williams LJ. Principal component analysis. Wiley Interdisciplinary Reviews: Computational Statistics. 2010;2(4):433–459. doi:10.1002/wics.101

28. James G, Witten D, Hastie T, Tibshirani R. An introduction to statistical learning. vol 112. Springer; 2013.

29. Castlight Health Inc. Bar Code Medication Administration. 2018. https://www.leapfroggroup.org/sites/default/files/Files/Leapfrog-Castlight_BCMA_Final.pdf

30. Pruitt ZM, Kazi S, Weir C, et al. A Systematic Review of Quantitative Methods for Evaluating Electronic Medication Administration Record and Bar-Coded Medication Administration Usability. Appl Clin Inform. Jan 2023;14(1):185–198. doi:10.1055/s-0043-1761435

31. Fox MP, MacLehose RF, Lash TL. Applying quantitative bias analysis to epidemiologic data. Springer; 2022.

32. Madaras-Kelly K, Jones M, Remington R, Hill N, Huttner B, Samore M. Development of an antibiotic spectrum score based on veterans affairs culture and susceptibility data for the purpose of measuring antibiotic de-escalation: a modified Delphi approach. Infection Control & Hospital Epidemiology. 2014;35(9):1103–1113.

33. Zhao L, Li Y, Jiang N, et al. Association of Blood Biochemical Indexes and Antibiotic Exposure With Severe Immune-related Adverse Events in Patients With Advanced Cancers Receiving PD-1 Inhibitors. J Immunother. May 1 2022;45(4):210–216. doi:10.1097/cji.0000000000000415

34. Finch WH, Finch MEH. Regularization methods for fitting linear models with small sample sizes: Fitting the lasso estimator using R. *Practical Assessment*, Research, and Evaluation. 2016;21(1):7.

35. Zhang C-H, Huang J. The sparsity and bias of the lasso selection in high-dimensional linear regression. 2008;

36. Bagherzadeh-Khiabani F, Ramezankhani A, Azizi F, Hadaegh F, Steyerberg EW, Khalili D. A tutorial on variable selection for clinical prediction models: feature selection methods in data mining could improve the results. Journal of Clinical Epidemiology. 2016/03/01/ 2016;71:76–85. 10.1016/j.jclinepi.2015.10.002

37. Sanchez-Pinto LN, Venable LR, Fahrenbach J, Churpek MM. Comparison of variable selection methods for clinical predictive modeling. International Journal of Medical Informatics. 2018/08/01/ 2018;116:10–17. 10.1016/j.ijmedinf.2018.05.006

38. Speiser JL, Miller ME, Tooze J, Ip E. A comparison of random forest variable selection methods for classification prediction modeling. Expert Systems with Applications. 2019/11/15/ 2019;134:93–101. 10.1016/j.eswa.2019.05.028

39. van der Ploeg T, Steyerberg EW. Feature selection and validated predictive performance in the domain of Legionella pneumophila: a comparative study. BMC Research Notes. 2016/03/08 2016;9(1):147. doi:10.1186/s13104-016-1945-2

40. Chowdhury MZI, Turin TC. Variable selection strategies and its importance in clinical prediction modelling. Fam Med Community Health. 2020;8(1):e000262. doi:10.1136/fmch-2019-000262

## References

1. Zhao, J., S. Murray, and J.J. LiPuma, Modeling the Impact of Antibiotic Exposure on Human Microbiota. Scientific Reports, 2014. 4(1): p. 4345.

2. Ben-Ami, R., et al., Antibiotic exposure as a risk factor for fluconazole-resistant Candida bloodstream infection. Antimicrobial agents and chemotherapy, 2012. 56(5): p. 2518–2523.

3. Song, J., et al., The Association Between the Frequency of Interruptions in Antibiotic Exposure and the Risk of Health Care-Associated Clostridiodes difficile Infection. Current Therapeutic Research, 2020. 93: p. 100600.

4. Stevens, V., et al., Differential risk of Clostridium difficile infection with proton pump inhibitor use by level of antibiotic exposure. Pharmacoepidemiol Drug Saf, 2011. 20(10): p. 1035–42.

5. Zhou, J.J., et al., Efficacy of bismuth-based quadruple therapy for eradication of Helicobacter pylori infection based on previous antibiotic exposure: A large-scale prospective, single-center clinical trial in China. Helicobacter, 2020. 25(6): p. e12755.

6. Huang, Q.M., M.A. Horn, and S.G. Ruan, Modeling the effect of antibiotic exposure on the transmission of methicillin-resistant Staphylococcus aureus in hospitals with environmental contamination. Math Biosci Eng, 2019. 16(5): p. 3641–3673.

7. Hung, Y.P., et al., Risk factors of fecal toxigenic or non-toxigenic Clostridium difficile colonization: impact of Toll-like receptor polymorphisms and prior antibiotic exposure. PLoS One, 2013. 8(7): p. e69577.

8. Korkmaz, H., et al., Effect of Antibiotic Exposure on Upper Respiratory Tract Bacterial Flora. Med Sci Monit, 2022. 28: p. e934931.

9. Caffrey, A.R., et al., Heterogeneity in the treatment of bloodstream infections identified from antibiotic exposure mapping. Pharmacoepidemiol Drug Saf, 2019. 28(5): p. 707–715.

10. Hui, C., et al., Previous antibiotic exposure and evolution of antibiotic resistance in mechanically ventilated patients with nosocomial infections. J Crit Care, 2013. 28(5): p. 728–34.

11. Schechner, V., et al., Antibiotic exposure and the risk of hospital-acquired diarrhoea and Clostridioides difficile infection: a cohort study. J Antimicrob Chemother, 2021. 76(8): p. 2182–2185.

12. Smith, J.T., et al., Quantifying the breadth of antibiotic exposure in sepsis and suspected infection using spectrum scores. Medicine (Baltimore), 2022. 101(41): p. e30245.

13. DiDiodato, G. and L. Fruchter, Antibiotic exposure and risk of community-associated Clostridium difficile infection: A self-controlled case series analysis. Am J Infect Control, 2019. 47(1): p. 9–12.

14. Song, J., et al., Temporal change of risk factors in hospital-acquired Clostridioides difficile infection using time-trend analysis. Infection Control & Hospital Epidemiology, 2020. 41(9): p. 1048–1057.

15. Rose, L., et al., Effects of aggregate and individual antibiotic exposure on vancomycin MICs for Staphylococcus aureus isolates recovered from pediatric patients. J Clin Microbiol, 2013. 51(9): p. 2837–42.

16. Maxfield, A.Z., et al., General antibiotic exposure is associated with increased risk of developing chronic rhinosinusitis. Laryngoscope, 2017. 127(2): p. 296–302.

17. Mertsalmi, T.H., E. Pekkonen, and F. Scheperjans, Antibiotic exposure and risk of Parkinson’s disease in Finland: A nationwide case-control study. Mov Disord, 2020. 35(3): p. 431–442.

18. Uldbjerg, C.S., et al., Antibiotic exposure during pregnancy and childhood asthma: a national birth cohort study investigating timing of exposure and mode of delivery. Arch Dis Child, 2021. 106(9): p. 888–894.

19. Fessas, P., et al., Early Antibiotic Exposure Is Not Detrimental to Therapeutic Effect from Immunotherapy in Hepatocellular Carcinoma. Liver Cancer, 2021. 10(6): p. 583–592.

20. Hoskin-Parr, L., et al., Antibiotic exposure in the first two years of life and development of asthma and other allergic diseases by 7.5 yr: a dose-dependent relationship. Pediatric Allergy and Immunology, 2013. 24(8): p. 762–771.

21. Morrell, S., et al., Antibiotic exposure within six months before systemic therapy was associated with lower cancer survival. J Clin Epidemiol, 2022. 147: p. 122–131.

22. Yang, S.I., et al., Effect of antibiotic use and mold exposure in infancy on allergic rhinitis in susceptible adolescents. Ann Allergy Asthma Immunol, 2014. 113(2): p. 160–165.e1.

23. Chapman, T.J., et al., Antibiotic Use and Vaccine Antibody Levels. Pediatrics, 2022. 149(5).

24. Horton, D.B., et al., Antibiotic exposure and juvenile idiopathic arthritis: a case–control study. Pediatrics, 2015. 136(2): p. e333–e343.

25. Sanyaolu, L.N., et al., Antibiotic exposure and the risk of colorectal adenoma and carcinoma: a systematic review and meta-analysis of observational studies. Colorectal Dis, 2020. 22(8): p. 858–870.

26. Horton, D.B., et al., Antibiotic exposure, infection, and the development of pediatric psoriasis: a nested case-control study. JAMA dermatology, 2016. 152(2): p. 191–199.

27. World Health Organization. Defined Daily Dose: Definition and general considerations. 2022 2/25/2023]; Available from: https://www.who.int/tools/atc-ddd-toolkit/about-ddd.

28. Benko, R., et al., Quantitative disparities in outpatient antibiotic exposure in a Hungarian county. J Antimicrob Chemother, 2008. 62(6): p. 1448–50.

29. Schechner, V., et al., Epidemiological interpretation of studies examining the effect of antibiotic usage on resistance. Clin Microbiol Rev, 2013. 26(2): p. 289–307.

30. Stanić Benić, M., et al., Metrics for quantifying antibiotic use in the hospital setting: results from a systematic review and international multidisciplinary consensus procedure. Journal of Antimicrobial Chemotherapy, 2018. 73(suppl_6): p. vi50–vi58.

31. Gerber, J.S., et al., Development and Application of an Antibiotic Spectrum Index for Benchmarking Antibiotic Selection Patterns Across Hospitals. Infection Control & Hospital Epidemiology, 2017. 38(8): p. 993–997.

32. Livorsi, D.J., et al., Antibiotic Stewardship Implementation and Antibiotic Use at Hospitals With and Without On-site Infectious Disease Specialists. Clin Infect Dis, 2021. 72(10): p. 1810–1817.

33. McCarthy, K.N., A. Hawke, and E.M. Dempsey, Antimicrobial stewardship in the neonatal unit reduces antibiotic exposure. Acta Paediatr, 2018. 107(10): p. 1716–1721.

34. National Health Safety Network, The NHSN Standardized Antimicrobial Administration Ratio (SAAR). 2023, CDC,.

35. Leung, V., et al., Metrics for evaluating antibiotic use and prescribing in outpatient settings. JAC-Antimicrobial Resistance, 2021. 3(3).

